# Explainable Machine Learning Predictions of Perceptual Sensitivity for Retinal Prostheses

**DOI:** 10.1101/2023.02.09.23285633

**Authors:** Galen Pogoncheff, Zuying Hu, Ariel Rokem, Michael Beyeler

## Abstract

To provide appropriate levels of stimulation, retinal prostheses must be calibrated to an individual’s perceptual thresholds (‘system fitting’), despite thresholds varying drastically across subjects, across electrodes within a subject, and over time. Although previous work has identified electrode-retina distance and impedance as key factors affecting thresholds, an accurate predictive model is still lacking. To address these challenges, we 1) fitted machine learning (ML) models to a large longitudinal dataset with the goal of predicting individual electrode thresholds and deactivation as a function of stimulus, electrode, and clinical parameters (‘predictors’) and 2) leveraged explainable artificial intelligence (XAI) to reveal which of these predictors were most important. Our models accounted for up to 77% of the perceptual threshold response variance and enabled predictions of whether an electrode was deactivated in a given trial with F1 and AUC scores of up to 0.740 and 0.913, respectively. Deactivation and threshold models identified novel predictors of perceptual sensitivity, including subject age, time since blindness onset, and electrode-fovea distance. Our results demonstrate that routinely collected clinical measures and a single session of system fitting might be sufficient to inform an XAI-based threshold prediction strategy, which may transform clinical practice in predicting visual outcomes.

## I. Introduction

To provide appropriate levels of stimulation, retinal prostheses must be calibrated to each subject’s amount of electrical current needed to elicit visual responses (*perceptual threshold*). In the case of the Argus II Retinal Prosthesis System (Vivani Medical, Emeryville, CA; formerly Second Sight Medical Products, Inc.) [1], this process is part of *system fitting*, where perceptual thresholds are used to populate subject-specific lookup tables that determine how the grayscale values of an image recorded by the external camera are translated into electrical stimuli. Current practices rely on a well-established adaptive up-down staircase procedure, which predicts perceptual thresholds with reasonable accuracy based on approximately 100 trials of a visual detection task [2].

However, perceptual thresholds have been shown to vary drastically not just across subjects, but also across electrodes within a subject as well as over time [2]–[5]. Thresholds often undergo sudden and large fluctuations that can last several weeks and cannot be explained by gradual changes in the implant-tissue interface [5]. This makes threshold estimation a major bottleneck in system fitting, as the procedure has to be performed for each electrode and repeated on a regular basis. With Argus II consisting of 60 electrodes, this procedure is time-consuming at best, but will quickly become infeasible as new devices are being developed that feature hundreds or even thousands of electrodes [6], [7].

Although previous work has identified electrode-retina distance and impedance as key factors affecting thresholds [2]– [4], the lack of accurate, automated threshold estimation frameworks to date may suggest that these two factors alone are far from comprehensive. Further complicating this matter, many of the clinical parameters presumed useful in threshold estimation are difficult or expensive to collect, are prone to measurement error, and may vary drastically between subjects. It is therefore paramount to know which parameters are worth collecting. An explainable predictive model fitted to a longitudinal dataset may help elucidate such parameters.

To address these challenges, we set out to develop explainable machine learning (ML) models that could:

- predict perceptual thresholds on individual electrodes as a function of stimulus, electrode, and clinical parameters (‘predictors’),
- infer deactivation of individual electrodes as a function of these parameters, and
- reveal which of these predictors were most important to perceptual thresholds and electrode deactivation.

Part of this work was previously presented in [8]. Other studies previously focused on linear models [3], [4], which provide easily interpretable model parameters, but are often not powerful enough to fit the data. Higher-complexity ML models may offer state-of-the-art prediction accuracy, but can be ‘black boxes’ whose predictions are inscrutable, and hence not actionable in a clinical setting. On the other hand, explainable artificial intelligence (XAI) relies on ML approaches that can explain why a certain prediction was made while maintaining high accuracy [9], [10]. Such models have the potential to transform clinical practice in predicting visual outcomes.

## II. Related Work

A handful of previous studies have investigated factors affecting perceptual thresholds in retinal prostheses [2]–[4], focusing on a range of stimulus (e.g., pulse polarity, pulse rate), electrode (e.g., area), and clinical (e.g., retinal thickness, position of the implant) parameters.

De Balthasar *et al*. [2] correlated perceptual thresholds with electrode impedance, electrode size, electrode-retina distance, and retinal thickness in six recipients of the Argus I epiretinal prosthesis. The study identified impedance and electrode-retina distance as critical factors for determining perceptual thresholds, but did not attempt to develop a predictive model.

Ahuja and colleagues [3] correlated perceptual thresholds with mean electrode-retina distance (averaged across all electrodes of a subject), the mean distance of electrodes from the fovea (‘electrode-fovea distance’), and the dark-adapted fullfield light threshold in 22 recipients of the Argus II epiretinal prosthesis. In addition to electrode-retina distance, the study identified the residual light threshold as a critical factor, but did not attempt to predict thresholds from these factors on individual electrodes.

Shivdasani and colleagues [4] correlated perceptual thresholds with a number of stimulus (return configuration, pulse polarity, pulse width, inter-phase gap, pulse rate), electrode (area and number of ganged electrodes), and clinical (retinal thickness, electrode-retina distance) parameters in three recipients of a suprachoroidal retinal prosthesis (Bionic Vision Australia). In addition to electrode-retina distance, the study identified the electrode configuration as important (lowest thresholds were achieved with a monopolar return, anodic-first stimulus polarity, short pulse widths with long inter-phase gaps, and high stimulation rates).

In summary, all three studies identified the distance of electrodes from the retinal surface (‘electrode-retina distance’) as a critical factor, with electrode size and retinal thickness having only a negligible effect on thresholds. However, these studies were either focused on a small number of subjects [2], [4] or were limited to predicting only the mean threshold across electrodes from a small number of factors [3]. A crossvalidated predictive model is still lacking.

It is worth noting that some of these parameters are more easily collected than others. For example, retinal thickness can only be inferred from optical coherence tomography (OCT) images, which is 1) difficult to collect as most retinal implant recipients present with nystagmus, and 2) error-prone due to electrodes casting shadows on the b-scan [3]. It is therefore paramount to know which of these parameters are worth collecting for the purpose of threshold prediction.

## II. Methods

### A. Dataset

We retrospectively analyzed a longitudinal dataset of 6,225 perceptual thresholds and electrode impedances measured on 673 electrodes in 13 Argus II patients (Table I; for demographic information see Table A.1 and [3], [11], [12]). The data was collected from 2007 − 2018 during 353 sessions conducted at 7 different implant centers located across the United States, the United Kingdom, France, and Switzerland. A subset of the data was previously collected as part of the Argus II Feasibility Protocol (Clinical Trial ID: NCT00407602). Our study, which did not involve human subjects research, was deemed exempt from institutional review board (IRB) approval by the IRB at the University of California, Santa Barbara.

**TABLE I:**
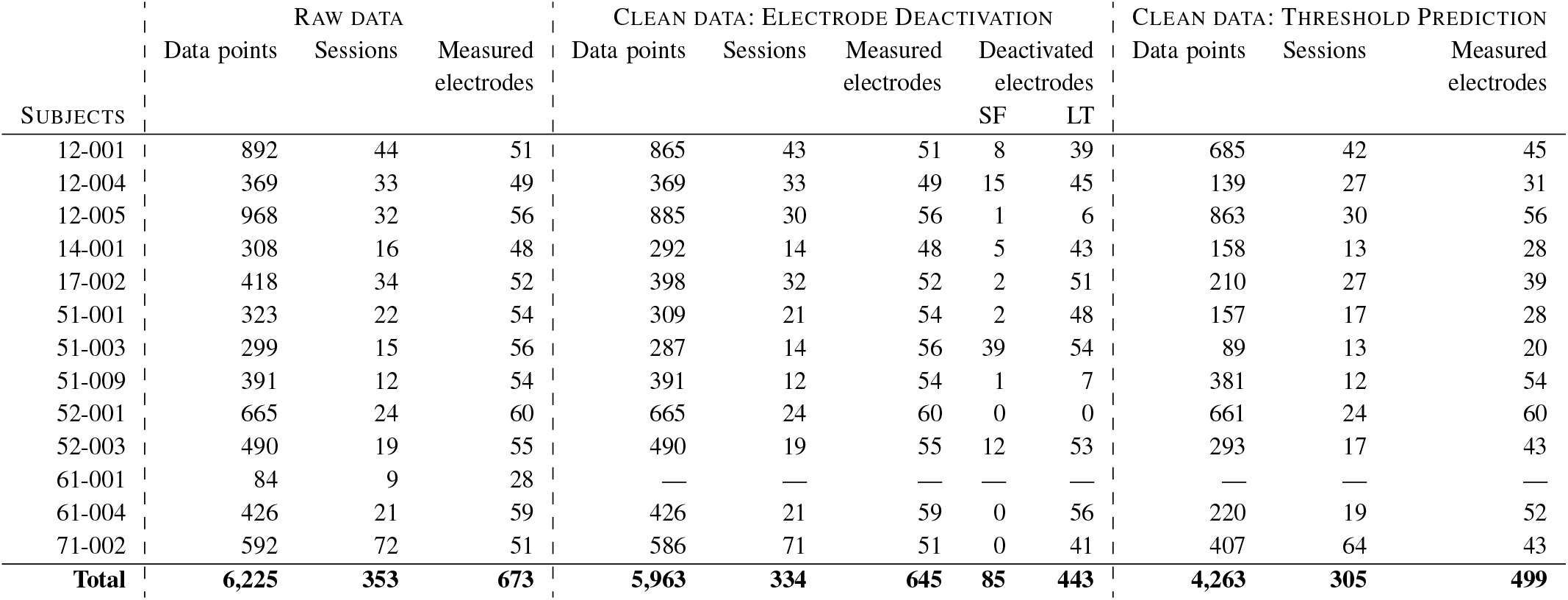
Summary of the Argus II dataset. SF: system fitting, LT: life time.

Data cleaning and preprocessing was performed in accordance with the two tasks central to our analyses (Subsection III-C) and is further detailed in Subsections III-E.1 and III-E.2. Given the limited amount of data available from Subject 61-001 after data cleaning, data from this subject was omitted from our analyses. For all 12 remaining subjects, threshold measurements were available for a majority of the 60 electrodes in the array, measured 12 − 72 times over the lifetime of the device. The manufacturer would often deactivate electrodes that were deemed nonfunctional, either because impedance measurements indicated an open or short circuit, or because no perceptual threshold below the charge density limit could be measured. Whereas only a handful electrodes were deactivated during system fitting (labeled ‘SF’ in Table I, *Clean data: Electrode Deactivation* column), most electrodes were at least temporarily deactivated over the lifetime of the device (labeled ‘LT’).

### B. Feature Engineering

To prepare the raw data for ML, we combined threshold and impedance values with clinical data sourced from the literature, and performed feature engineering. Given our goal of discovering the core predictors of perceptual sensitivity to stimulation in patients with Argus II implants, all engineered features used in the modeling proposed in this work were domain-specific and constructed to be of direct, practical use to clinicians and prosthetic vision researchers.

The resulting feature correlation matrix is shown in Fig. 1, with each feature described in Table II and in more detail below. As some parameters are more easily collected than others, a major goal of this work was to identify which of the available parameters are worth collecting for the purpose of predicting perceptual thresholds. We therefore split the available features into three different categories:

**Fig. 1:**
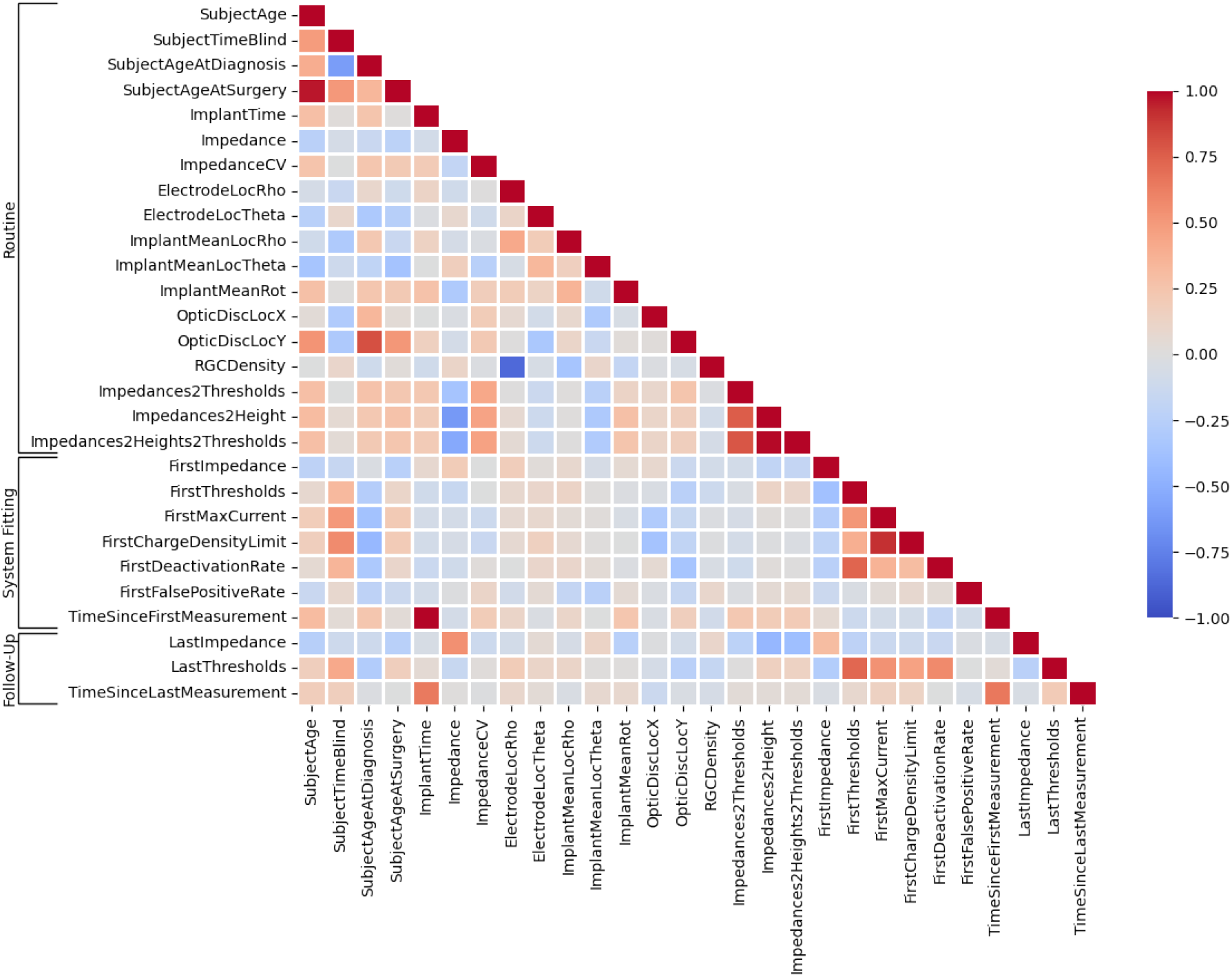
Feature correlation heat map.

**TABLE II:** Features (measured and engineered) for perceptual sensitivity prediction.

|  | Feature | Description | Precision |
| --- | --- | --- | --- |
| Routine | SubjectAge | Subject age | days |
|  | SubjectTimeBlind | Estimated time since onset of blindness | days |
|  | SubjectAgeAtDiagnosis | Subject age at first retinitis pigmentosa (RP) diagnosis | years |
|  | SubjectAgeAtSurgery | Subject age at time of implant surgery | years |
|  | ImplantTime | Time since implant surgery | days |
| | Impedance | Manufacturer-provided impedance reading at each electrode | kΩ |
|  | ImpedanceCV | Coefficient of variation for impedance values | float |
| | ElectrodeLoc{Rho \| Theta} | Electrode-fovea distance | $\mu\text{m}$ \| rad |
| | ImplantLoc{Rho \| Theta \| Rot} | Mean electrode-fovea distance | $\mu\text{m}$ \| rad \| rad |
|  | OpticDiscLoc{X \| Y} | Optic disc location | deg |
|  | RGCDensity | Retinal ganglion cell (RGC) density [13] | RGC/deg<sup>2</sup> |
| | Impedances2Thresholds | Threshold estimated from impedance values [2] | $\mu\text{A}$ |
| | Impedances2Height | Electrode-retina distance estimated from impedance values [2] | $\mu\text{m}$ |
| | Impedances2Height2Thresholds | Threshold estimated from estimated electrode-retina distances [2] | $\mu\text{A}$ |
| System Fitting | FirstImpedance | Impedance reading at system fitting for each electrode | kΩ |
| | FirstThreshold | Perceptual stimulus threshold at system fitting for each electrode | $\mu\text{A}$ |
| | FirstMaxCurrent | Largest allowable current at system fitting | $\mu\text{A}$ |
|  | FirstChargeDensityLimit | Charge density limit at time of system fitting | mCcm<sup>-2</sup> |
|  | FirstDeactivationRate | Fraction of electrodes deactivated at system fitting | float |
|  | FirstFalsePositiveRate | False positive phosphene perception rate during system fitting | float |
|  | TimeSinceFirstMeasurement | Elapsed time since system fitting | days |
| Follow-Up | LastImpedance | Impedance reading from previous session for each electrode | kΩ |
| | LastThreshold | Perceptual stimulus threshold from previous session for each electrode | $\mu\text{A}$ |
|  | TimeSinceLastMeasurement | Time elapsed previous electrode threshold measurement | days |

#### 1) Routinely Collected Data

We sourced public information about patient history (e.g., age at blindness diagnosis, age at implant surgery) from previous studies [3], [11], [12]. Surgery dates were obtained from Second Sight, and where not known exactly, were triangulated from the known dates of the earliest available impedance measurements and the 3-month followup exam. Knowing surgery dates and the subject’s age at that time allowed us to estimate the birth year for each subject (±1 year). Based on the above information and the dates of each testing session we were therefore able to calculate for each testing session: i) time since surgery, ii) time since diagnosis, and iii) subject age at each session.

Implant placement and optic disc location were estimated using retinal fundus images obtained either 12 months, 24 months, or 36 months after surgery. Since we did not have access to fundus images from each session, we assumed that the implant did not move over time [14]. We used a procedure described in [15] and the pulse2percept software [16] to perform image registration and extract the location of the implant and each electrode with respect to the fovea (Fig. 2). With these locations, retinal ganglion cell (RGC) density was estimated following the methods of Curcio and Allen in [13].

**Fig. 2:**
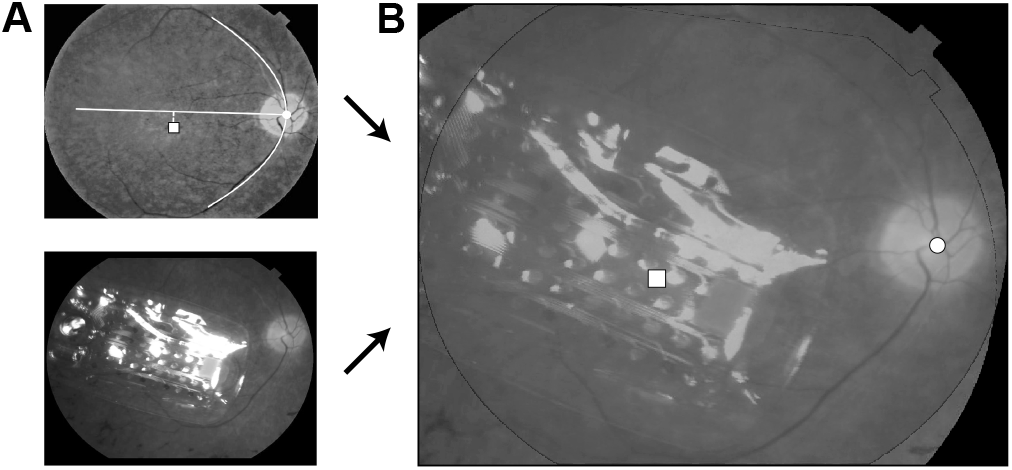
The location and orientation of each subject’s implant was estimated by combining their postsurgical fundus photograph (*A*, bottom) with a baseline presurgical image in which the fovea had been identified (*A*, top) to produce a registered image (*B*; □: foveal pit, : optic disc). The horizontal raphe (*A*, white line) was approximated by fitting a parabola to the main vascular arcade and finding the tangent to the parabola inflexion point (adapted under CC-BY from [15]).

As mentioned above, previous work identified electrode-retina distance as a key factor affecting thresholds [2]– [4]. Unfortunately, we did not have access to OCT images for all subjects. Instead we followed de Balthasar *et al*. [2] to estimate electrode-retina distance from the available impedance measurements (‘Impedance2Height’ in Table II). Additional relationships between impedance, electrode-retina distance, and thresholds proposed by [2] were used to obtain surrogate threshold estimates (‘Impedances2Thresholds’ and ‘Impedances2Height2Thresholds’) from routinely collected measurements of impedance.

Additional predictors relevant to perceptual sensitivity outcomes that were available but omitted from this work include a categorical variable specifying the clinic at which the Argus II implant operation was performed for each subject, an indicator of how many Argus II implant operations had been performed at the site, a binary variable specifying whether the device was implanted in the subject’s left or right eye, and the sex of the subject. Although visual outcomes may also depend on surgical precision (as complications during implantation could exacerbate fibrosis), predictors relevant to the surgical center were not considered in our study in an effort to focus our analyses on factors relevant to the subject and implant. Additionally, in concern of model generalization, features relevant to implant eye and subject sex were removed, because only two out of twelve subjects were female and only one subject had the implant in their left eye.

#### 2) System Fitting

Soon after system activation, patients undergo a system fitting procedure during which electrode impedances and perceptual thresholds are measured on all 60 electrodes. These values are then used to set several system parameters, such as the charge density limit and the largest allowable current. By default, charge density limits are set to 0.35 mC cm^−2^ per phase for everyday use, and to 1 mC cm^−2^ for lab use, but can be reduced for electrodes that are particularly sensitive. Analogously, stimulating currents are limited to 1 mA per default, but can be reduced for sensitive electrodes. Electrodes whose impedance value indicate either a short or open circuit are immediately deactivated.

Features derived from measurements obtained during system fitting (Table II, middle section) were only used as features for the training data, never as labels that the algorithm was supposed to predict. Several additional features, such as the fraction of deactivated electrodes and the false positive rate during threshold measurements (where the stimulating current was zero but the patient reported seeing a phosphene), were engineered using the observations obtained during system fitting. Finally, for each measurement sample in the dataset, we calculated the time that had passed since system fitting.

#### 3) Follow-Up Examinations

Patients participating in the Argus II Feasibility Protocol regularly visited their eye clinic for follow-up exams. We wondered how useful these more recent threshold and impedance measurements were for predicting perceptual sensitivity (Table II, bottom section). Since the time between sessions varied, we also calculated the time that had passed since the last examination.

### C. Prediction Tasks

We studied predictors of perceptual sensitivity in the context of two prediction tasks:

- *Threshold prediction:* a regression task in which the goal of the ML model was to predict the perceptual threshold on a given electrode at a specific point in time.
- *Electrode deactivation:* a binary classification task in which the goal of the ML model was to correctly predict whether or not an electrode was deactivated during at a specific point in time.

Given the available data, these two tasks enabled us to directly study the impact of each feature on quantitative measures of perceptual sensitivity by means of XAI. While we expected that a subset of the predictors most important to the ML models trained for each of these tasks would be shared, we hypothesized that the models developed for the regression task would better reveal predictors that contribute to finer-granularity fluctuations in perceptual sensitivity.

### D. Explainable Machine Learning Models

As in many neural engineering applications where ML models are used in decision-making processes, it is critical that the predictions made by the model are explainable. Specific to perceptual outcome prediction, we aimed to develop models that can inform clinicians of the most relevant parameters to collect and how such parameters may be used to automate stimulus threshold parameterization. In pursuit of these goals, we considered both linear and nonlinear ML models.

We leveraged logistic regression (LR) with L1 and L2 loss constraints for electrode deactivation and Elastic Net (EN) linear models for threshold regression. We chose these models both for their simplicity, explainability, and robustness to correlated features afforded by L1 loss constraints. Meanwhile, in addition to the regularization that results from L1 and L2 loss, we expected that these low-variance models would elucidate the perceptual outcome predictors that were most generalizable across subjects in our logitudinal dataset.

In addition, we used gradient boosting models (specifically, XGBoost models) in our non-linear modeling analysis. Unlike linear models, whose output is based on a linear, weighted combination of input feature values, XGBoost models are based on ensembles of decision trees [17] defined on multiple subsets of dataset features, allowing them to capture highly nonlinear patterns in the underlying data. Such models have enabled state-of-the-art results in a variety of practical tasks with heterogeneous features and complex dependencies.

In modeling our data, we aimed to discover the most salient features relevant to perceptual sensitivity, whether these relationships were linear or not. Furthermore, the two model types were used to establish benchmark results for electrode deactivation and perceptual threshold prediction. To apply these models to our longitudinal data, we assumed that each testing session was independent. We thus treated timestamp-related data (e.g., ‘SubjectTimePostOp’, ‘SubjectTimeBlind’) as additional feature attributes.

To determine the relative importance of the different features for each model’s predictions, we adopted the use of SHapley Additive exPlanations (SHAP) [18]. SHAP is a feature attribution technique based on the game-theoretically optimal Shapley values, which determine how to fairly distribute a ‘payout’ (i.e., the prediction) among model parameters. As SHAP analysis is model-agnostic, it is applicable to linear and non-linear models in both electrode deactivation and threshold prediction tasks. Although these models can be explained through their fitted parameters, SHAP enables a more direct comparison of the predictive behavior between models with varied parameters and assumptions about the underlying data. Additionally, SHAP values offer insight into a model’s decision at the granularity of a single test sample.

In the analyses that follow, LR and EN models were implemented using scikit-learn’s LogisticRegression and ElasticNet APIs (v1.1.2). XGBoost models were implemented using the XGBClassifier and XGBRegressor APIs of the XGBoost package (v1.6.2). Bayesian hyperparameter optimization was performed using the scikit-optimize package (v0.9.0). The code to run the models and generate the figures can be found at https://github.com/bionicvisionlab/2023-ArgusThresholds.

### E. Model Evaluation and Comparison

A significant challenge in developing ML models for biological data is the inherent inter-subject variability of such data. It is not uncommon for a data distribution from one subject to be divergent from the data distribution of another (Fig. A.1). To estimate the performance of our proposed electrode deactivation and threshold prediction models on data from unobserved subjects, we therefore employed a leave-onesubject-out (LOSO) analysis as follows: for each of the twelve subjects, we instantiated a new model, trained the model on data from the remaining eleven subjects, and finally generated predictions exclusively for the data of the held-out test-subject.

To allow for a fair comparison between models, we performed Bayesian hyperparameter optimization in a nested LOSO cross-validation loop. During this hyperparameter optimization process, the posterior probability distribution of an objective score (F1-score in the case of electrode deactivation and *R*^2^ in the case of threshold prediction) was estimated, given the model’s set of hyperparameters. We then selected the hyperparameters expected to maximize the objective score on the validation data and re-fit the model to the entire training dataset prior to testing. Hyperparameters optimized for the LR classification model included ‘C’ and ‘l1 ratio’, controlling the total regularization strength and the portion of this regularization contributed by L1 loss. Similarly, hyperparameters (‘alpha’ and ‘l1 ratio’) were optimized for the EN regression model. For both the XGBClassifier (XGB-C) and XGBRegressor (XGB-R) models, we tuned the number of estimators in the ensemble (‘n estimators’), the max depth of each decision tree (‘max depth’), partition criteria ‘min child weight’ and ‘gamma’, and L1 and L2 regularization terms (‘reg alpha’ and ‘reg lambda’, respectively).

We evaluated model performance when trained with the three subsets of predictors described above (Table II): ‘Routine’ predictors, a set composed of the 18 predictors derived from routinely collected clinical data; ‘System Fitting’ predictors, a subset which contained the predictors of the ‘Routine’ subset as well as the 7 predictors derived from measurements obtained during system fitting; and ‘Follow-Up’ predictors, which contained the 25 predictors from the ‘System Fitting’ subset in addition to the 3 follow-up trial predictors. For ‘System Fitting’ and ‘Follow-Up’, the first measurement from each electrode was removed from the dataset to prevent leaking ground-truth data into the feature vectors.

#### 1) Threshold Prediction

During threshold regression, models inferred a real-valued perceptual threshold for each electrode. In this task, recordings associated with electrodes that were deactivated in the session were removed, as no sensible threshold current could be assigned to the deactivated electrode in this case. As in the electrode deactivation task, we removed trials with missing data or impedance readings of 0 kΩ and normalized feature values. Furthermore, as perceptual threshold estimation is a high-variance procedure, it was not uncommon to observe within-session variability of the threshold for a given electrode or for subjects to report the presence of a phosphene in the absence of stimulation (‘catch trials’). To account for such sources of noise in threshold prediction, we discarded outlier samples using an automated, statistical method based on Chebyshev’s data distribution tail bounds [19]. Model performance was quantitatively analyzed with the adjusted coefficient of determination 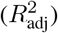 and a variant of the fraction of explainable variance explained (FEVE) [20].

FEVE ∈ (−∞, 1] offers a quantitative measure of explainable variance, like *R*^2^, while also accounting for variability in measurements of the dependent variable of interest (i.e., perceptual threshold) influenced by uncontrolled factors during measurement (e.g., perceptual lapses and false positive perceptions). FEVE was computed as follows:

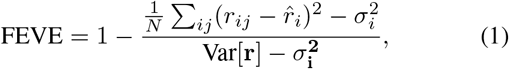

where *r*_*ij*_ was the *j*-th ground-truth perceptual threshold measurement for electrode *i* in a single recording session, 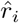 was the predicted perceptual threshold for this electrode during the session, *N* was the total number of perceptual threshold observations, Var[**r**] was the variance of all perceptual threshold measurements **r**, and 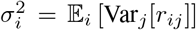 was the expected variance in perceptual threshold measurements for stimuli presented at each electrode of each subject.

Unlike in [20], however, in which identical stimuli are repeatedly presented in multiple trials, it was uncommon for repeated threshold measurements to be made for the same electrode on any given day. For this reason, we derived a similar measure, FEVE_*d*_, which assumed that the perceptual threshold of a given electrode remained constant over a period of *d* days (i.e., responses **r** within *d* days of one another were treated as repeated measurements for the given set of electrodes). Note that FEVE_0_ FEVE from Eq. 1.

Adding to the difficulty of modeling data with a significant degree of inter-subject variability, it is not uncommon to observe diverging perceptual threshold distributions between subjects (Fig. A.1). To account for these differences when modeling perceptual thresholds with ‘System Fitting’ and ‘Follow-Up’ data subsets, we scaled each threshold measurement according to the first threshold measured for the electrodes of each subject instead of directly predicting perceptual thresholds. This transformation implies that the models fitted to ‘System Fitting’ and ‘Follow-Up’ feature sets learned to predict *changes* in perceptual thresholds, relative to system fitting measurements. We found that this enabled better model generalization in LOSO threshold prediction. Note that this was not possible when modeling with the ‘Routine’ feature subset, however, since we assumed that no previous threshold measurements were available in this scenario.

#### 2) Electrode Deactivation

For the task of electrode deactivation, each electrode was assigned class 1 if it was deactivated in the given recording session and 0 otherwise. Trials with missing values or invalid impedance readings (i.e., 0 kΩ) were removed from the dataset prior to model fitting and analysis. In each LOSO iteration, the synthetic minority oversampling technique (SMOTE) [21] was applied to the training dataset to balance the number of samples from each class. All feature values were normalized according to the distribution of the training data to have zero mean and unit standard deviation.

## IV. Results

### A. Factors Affecting Perceptual Sensitivity

Fig. 3 shows the Spearman correlation coefficient for the 28 predictors, ordered by correlation magnitude. Aside from historical threshold measurements (i.e., ‘LastThresholds’ and ‘FirstThresholds’) which correlated strongest with perceptual sensitivity measures in future sessions, measures of impedance and estimates of electrode-retina distance (‘Impedances2Height’) and threshold from these impedance measurements (‘Impedances2Thresholds’) were among the predictors that had highest correlation with perceptual sensitivity. In terms of demographic factors, time since blindness onset (‘SubjectTimeBlind’) and time since implantation (‘ImplantTime’) had the highest correlations.

**Fig. 3:**
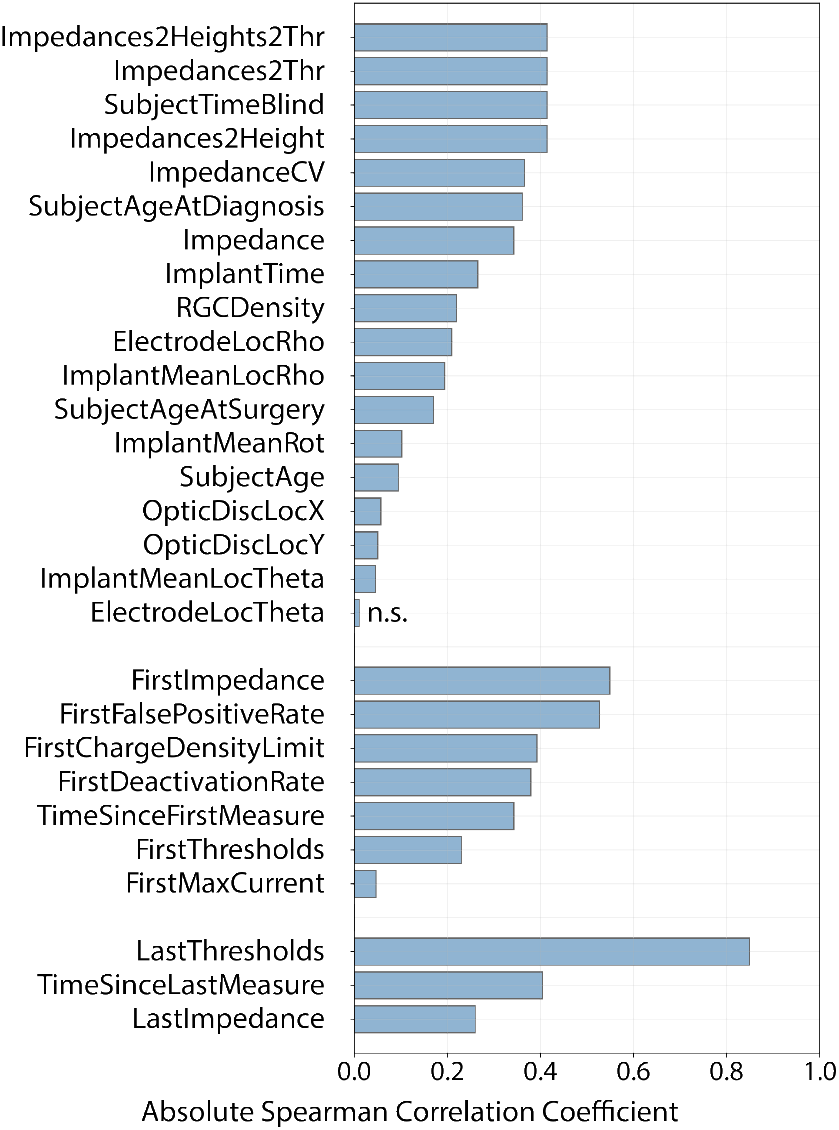
Magnitude of Spearman correlation coefficient between each predictor and perceptual thresholds, grouped by feature set (top: ‘Routine’, middle: ‘System Fitting’, bottom: ‘FollowUp’) and sorted by correlation magnitude. All predictors had *p <* .05 except for ‘ElectrodeLocTheta’ (labeled n.s.).

Fig. 4 shows the correlation between perceptual thresholds and the 28 predictors. Immediately observable in this figure is the high degree of variability among perceptual threshold measurements for any given predictor and the linear correlation between many of these predictors and perceptual sensitivity. When fitting linear regression coefficients between these predictors and their accompanying threshold measurements, the line of best fit for all but two predictors (labeled ‘ImplantMeanLocTheta’ and ‘FirstFalsePositiveRate’ in Fig. 4, respectively) had a significantly non-zero slope (*p <* 0.05). It is worth noting, however, that such trends were not always consistent across all 12 subjects.

**Fig. 4:**
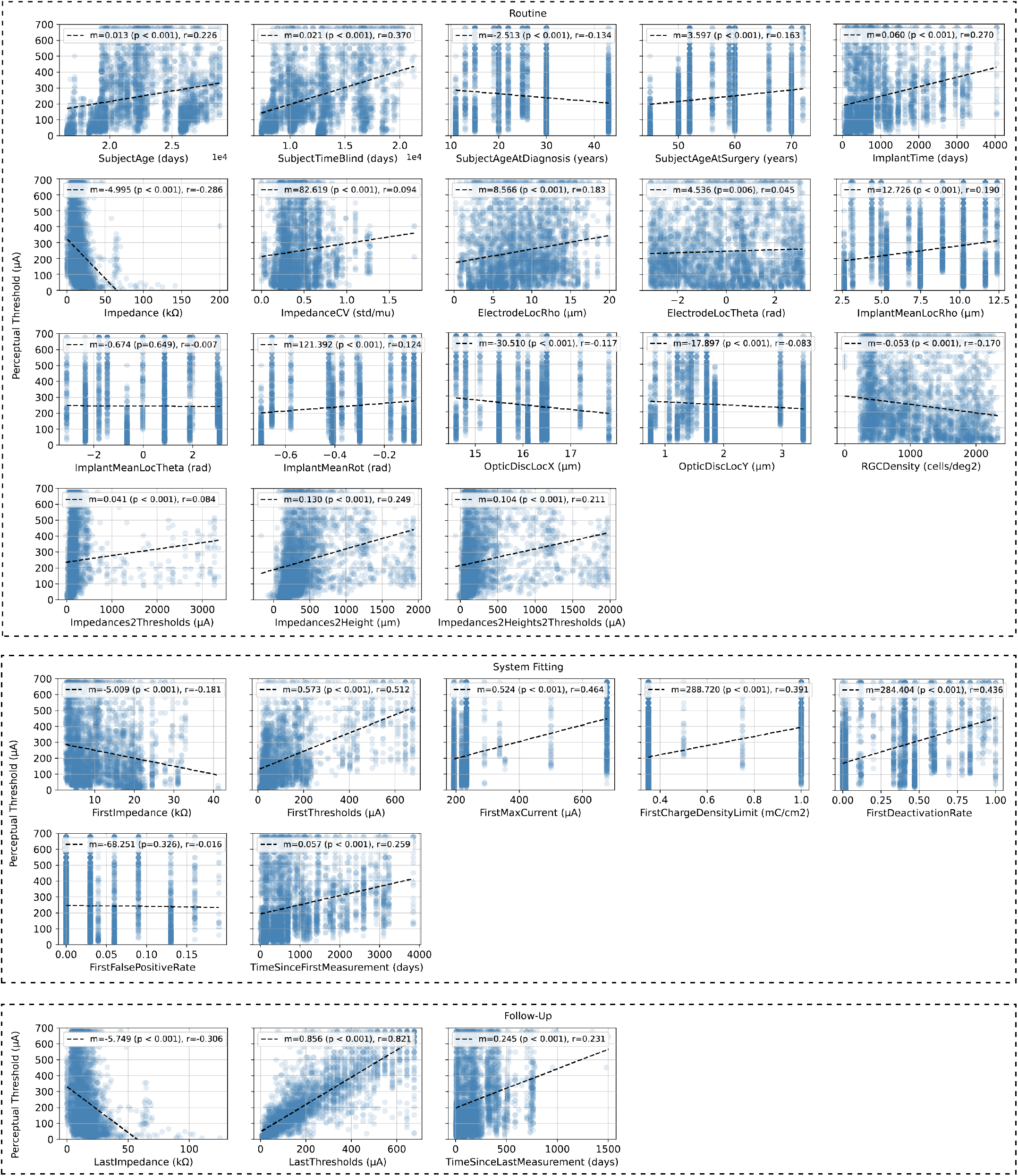
Predictor measurements plotted against perceptual stimulation thresholds and the associated linear-least squares regression line. The corresponding slopes (*m*), *p* values (associated with a null hypothesis that the slope of the linear regression line is zero), and Pearson Correlation Coefficients (*r*) are provided in the legend of each plot. Subplots are grouped by feature set (‘Routine’, ‘System Fitting’, or ‘Follow-Up’) and ordered by their listing in Table II.

Whereas previous studies were able to demonstrate that thresholds are negatively correlated with impedance readings (labeled ‘Impedance’ in Fig. 4) and positively correlated with electrode-retina distance (‘Impedances2Height’) [2]–[4], our data also highlights correlations with demographic factors. Most notably, thresholds tended to increase with subject age (‘SubjectAge’), subject age at implantation (‘SubjectAgeAtSurgery’), and time since blindness onset (‘SubjectTimeBlind’). Not surprisingly, thresholds also tended to increase with time since implantation (‘ImplantTime’), which is consistent with other studies [5]. In terms of neuroanatomical parameters, thresholds were positively correlated with electrode-fovea distance (‘ElectrodeLocRho’) and negatively correlated with ganglion cell density (‘RGCDensity’), which is a nonlinear function of electrode-fovea distance.

Thresholds over time were also strongly correlated with different measures typically obtained during system fitting, such as impedance and threshold readings (‘FirstImpedance’ and ‘FirstThreshold’, respectively) as well as established upper bounds on the allowed current amplitude (‘FirstMaxCurrent’) and charge density (‘FirstChargeDensityLimit’). Correlations were similar for predictors obtained during follow-up exams.

### B. Threshold Prediction

Table III shows aggregated LOSO threshold prediction results observed when modeling ‘Routine’, ‘System Fitting’, and ‘Follow-Up’ datasets with EN and XGB-R models. Results are presented for FEVE ≡ FEVE_0_ and FEVE_180_, where the latter assumed that perceptual thresholds remained stable over a period of 180 days (see Subsection III-E). Note that in some cases FEVE values were even more negative than 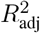, because of the subtraction of 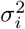 in Eq. 1. Perceptual threshold estimates compared to ground-truth for all 12 subjects can be found in Figs. B.2–B.3.

**TABLE III:** Leave-One-Subject-Out Perceptual Threshold Regression Results. Each metric is evaluated over an aggregated test set. Per-subject metric means and standard deviations are reported in parentheses.

| | Method | $R_{\text{adj}}^2$ | | FEVE | | FEVE<sub>180</sub> | |
| --- | --- | --- | --- | --- | --- | --- | --- |
| Routine | ElasticNet | -1.752 | (-3.034 ± 4.811) | -1.828 | (-2.930 ± 4.892) | -1.925 | (-3.209 ± 5.216) |
|  | XGB-Regressor | **-0.230** | (-1.545 ± 2.417) | **-0.236** | (-1.444 ± 2.497) | **-0.249** | (-1.610 ± 2.778) |
| System Fitting | ElasticNet | 0.133 | (-0.767 ± 1.780) | 0.140 | (-0.885 ± 2.620) | 0.145 | (-0.785 ± 2.434) |
|  | XGB-Regressor | **0.385** | (-0.056 ± 0.416) | **0.391** | (0.041 ± 0.444) | **0.405** | (0.080 ± 0.440) |
| Follow-Up | ElasticNet | **0.765** | (0.378 ± 0.401) | **0.772** | (0.455 ± 0.386) | **0.799** | (0.536 ± 0.392) |
|  | XGB-Regressor | 0.725 | (0.389 ± 0.315) | 0.732 | (0.490 ± 0.237) | 0.758 | (0.565 ± 0.251) |

Both EN and XGB-R models failed to yield accurate perceptual threshold predictions when relying solely on routinely collected data, as indicated by negative 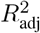 and FEVE values. Although many of these routine predictors correlated with perceptual thresholds, the regression results suggest that they alone do not carry sufficient information to predict perceptual thresholds over time.

Upon the introduction of measurements recorded during each subject’s system fitting session, the predictive power of the XGB-R increased notably 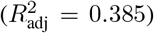, more so than the linear model 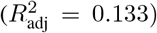. Finally, when considering all collected and engineered predictors in our models (i.e., the ‘Follow-Up’ feature subset), both EN and XGB-R models achieved accurate threshold predictions, explaining more than 70% of the variance in the data.

Post-hoc SHAP analysis (Fig. 5, top two rows) revealed that among all ‘System Fitting’ features, the XGB-R predictions were most influenced by initial threshold measurements from system fitting, the time elapsed since implant surgery, and the time elapsed since system fitting (‘FirstThresholds’, ‘ImplantTime’, and ‘TimeSinceFirstMeasurement’, respectively). In these plots, each data point is associated with a threshold prediction from the held-out cross-validation fold (test set). SHAP values indicate each feature’s contribution to the model’s prediction, with high SHAP values pushing the model towards predicting high thresholds, and low SHAP values pushing the model towards predicting low thresholds. As previously observed in [3], [4], measures of impedance (from both the current session and from system fitting) were also instrumental in this model’s decision process.

**Fig. 5:**
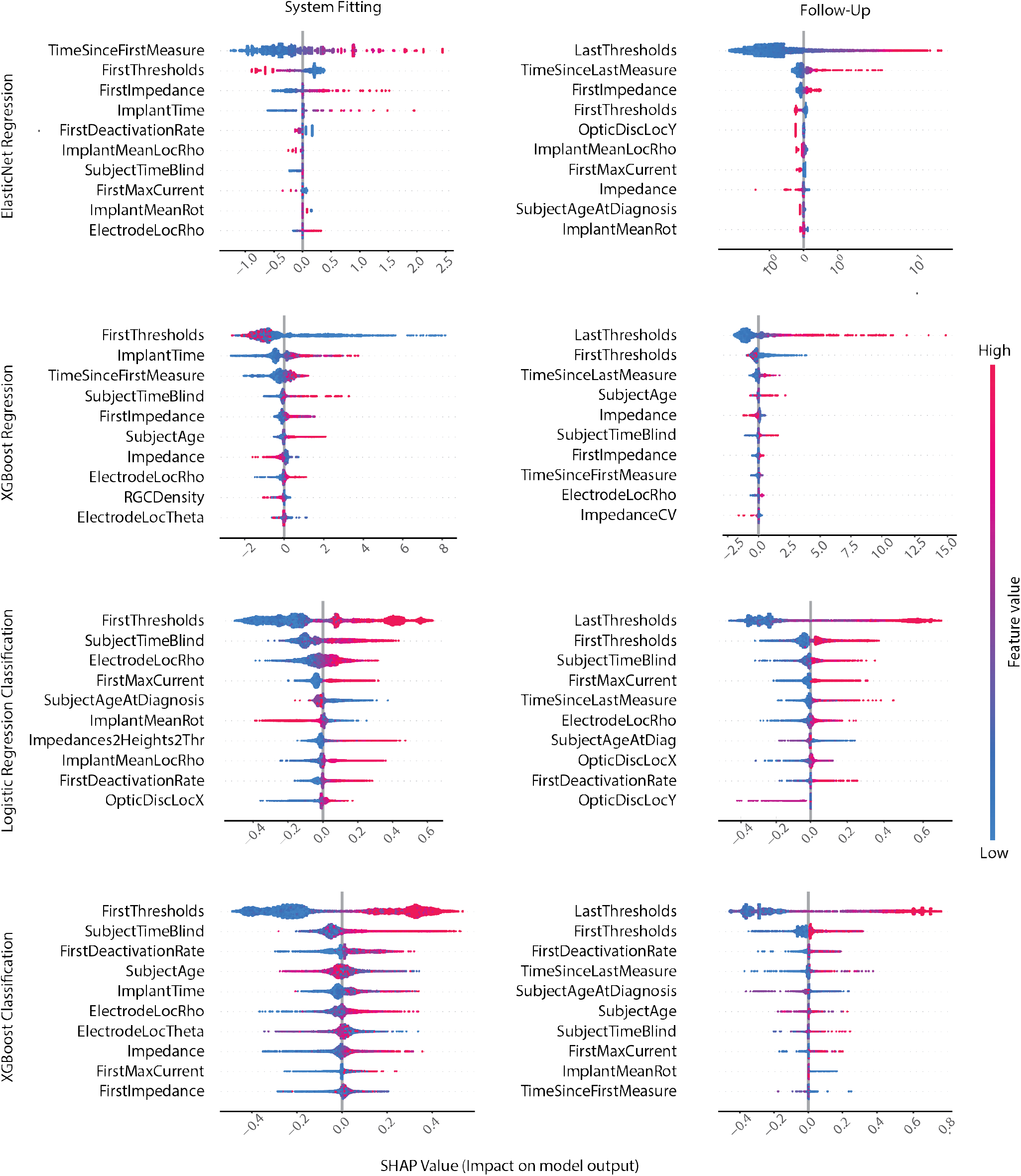
Force plot of SHapley Additive exPlanations (SHAP) values for the ten predictors that had greatest impact on threshold prediction (top two rows) and electrode deactivation inference (bottom two rows). Each data point is associated with a prediction from the held-out test set. SHAP values indicate each feature’s contribution to the model’s decision. For threshold prediction, predictors with high SHAP values influenced the model towards predicting greater threshold values while those smaller SHAP values encouraged the model to predict relatively lower threshold values. In the case of electrode deactivation, positive SHAP values indicate that the feature pushed the model towards predicting deactivation while negative values reflect that the feature pushed the model away from predicting deactivation. Colors indicate predictor values. Results are shown for ‘System Fitting’ and ‘Follow-Up’ feature splits (see Table II).

The two models, given their inherently different assumptions about the relationships between the predictors and perceptual thresholds, yield different insights into the most impacting predictors of perceptual threshold. The most important ‘Follow-Up’ predictor for both model types was the threshold measurement from the previous visit (‘LastThresholds’). This is unsurprising, considering its strong correlation with the electrode’s current perceptual threshold. Greater values for ‘LastThresholds’ tended to influence the model towards predicting a larger perceptual threshold sensitivity. Additional predictors shared between EN and XGB-R models included ‘FirstThresholds’, ‘FirstImpedance’, ‘TimeSinceLastMeasurement’, and ‘Impedance’. Of the top five predictors influencing the predictions of the EN model, the remaining four were ‘TimeSinceLastMeasurement’, ‘FirstImpedance’, ‘FirstThresholds’, and ‘OpticDiscLocY’. More time elapsed since a subject’s previous visit often biased the model towards predicting an increased perceptual threshold. Similarly, high impedance values measured at system fitting tended to lead towards the prediction of a higher perceptual threshold. Interestingly, a high initial threshold (measured during system fitting) led to decreased threshold predictions. Exclusively meaningful to the XGB-R model, advanced age (accounted for in the predictor ‘SubjectAge’) often led to higher threshold predictions in the XGB-R model.

Despite the improved generalization achieved by prediction of a scaled threshold value (as opposed to the exact perceptual threshold current; see Section III-E) in the case of modeling with ‘System Fitting’ and ‘Follow-Up’ features, impractical predictions were observed in rare occasions. Notably, an unbounded regression model output permitted the prediction of a negative perceptual threshold, but this was observed on only four out of 6,225 occasions.

Overall, these models offer two unique perspectives on the predictors most salient to thresholds in Argus II implants.

### C. Predicting Electrode Deactivation

Table IV shows aggregated LOSO classification results observed when modeling ‘Routine’, ‘System Fitting’, and ‘Follow-Up’ datasets with LR and XGB-C models.

**TABLE IV:** Results for Leave-One-Subject-Out (LOSO) Electrode Deactivation Classification. Each metric is evaluated over an aggregated held-out test set. Means and standard deviations across subjects are reported in parentheses. Subjects with no deactivated electrodes were excluded from the mean and standard deviation aggregation reported in parentheses.

|  | Method | Precision | Recall | F1 | AUC |
| --- | --- | --- | --- | --- | --- |
| Routine | Logistic regression | **0.237** (0.248 ± 0.259) | 0.392 (0.519 ± 0.434) | **0.295** (0.260 ± 0.288) | 0.466 (0.626 ± 0.140) |
| | XGB-Classifer | 0.219 (0.376 ± 0.337) | **0.408** (0.512 ± 0.360) | 0.285 (0.324 ± 0.282) | **0.475** (0.645 ± 0.145) |
| System Fitting | Logistic regression | **0.558** (0.447 ± 0.289) | 0.746 (0.625 ± 0.383) | **0.639** (0.502 ± 0.310) | **0.813** (0.733 ± 0.125) |
| | XGB-Classifer | 0.500 (0.462 ± 0.243) | **0.765** (0.663 ± 0.289) | 0.605 (0.530 ± 0.261) | 0.809 (0.693 ± 0.152) |
| Follow-up | Logistic regression | **0.648** (0.545 ± 0.271) | 0.843 (0.692 ± 0.351) | 0.733 (0.602 ± 0.295) | 0.913 (0.816 ± 0.133) |
| | XGB-Classifer | 0.646 (0.536 ± 0.265) | **0.867** (0.712 ± 0.344) | **0.740** (0.609 ± 0.295) | **0.913** (0.758 ± 0.171) |

LR and XGB-C models performed similarly when predicting electrode deactivation using each subset of predictors. Interestingly, without ever measuring perceptual thresholds (‘Routine’), both LR and XGB-C models were able to predict future electrode deactivations with an area under the ROC curve (AUC) of 0.47. In this case, the model decisions relied on initial impedance measurements as well as clinical information about the subject’s age, time since blindness onset, and time since device implantation. By incorporating threshold measurements as well as other parameters typically collected during system fitting, performance increased to an AUC of 0.8. When additional measurements from follow-up examinations were included, both models reached a peak AUC of 0.913.

The SHAP values for the ten most important features in the dataset, according to the ‘System Fitting’ and ‘FollowUp’ models, are shown in Fig. 5 (bottom two rows). In these plots, each data point is a prediction of electrode deactivation from the held-out cross-validation fold (test set). SHAP values indicate each feature’s contribution to the model’s decision, with positive values indicating that a feature pushed the model towards predicting deactivation, and negative values pushing the model away from predicting deactivation. Of all the routine clinical measures, subject age at diagnosis and the amount of time the subject has been blind were observed as the most important features for electrode deactivation inference. Regardless, low electrode deactivation classification performance was attained with LR and XGB-C models when relying exclusively on routine clinical measures.

A significant improvement in predictive performance was observed upon the introduction of threshold measurements and related electrode-specific settings typically obtained during system fitting. Specifically, initial thresholds and the proportion of electrodes deactivated during system fitting were important predictors for both LR and XGB-C models. Electrodes with greater initial threshold were more likely to be deactivated in the future. Similarly, higher proportions of deactivated electrodes during system fitting tended to influence the model towards predicting electrode deactivation for that subject. Higher electrode-fovea distances (‘ElectrodeLocRho’) biased both models towards predicting deactivation.

Not surprisingly, when data from follow-up examinations was considered as well, the most recently obtained threshold measurement proved to be the most important predictor. This is consistent with the finding that threshold measurements often go through large fluctuations over time [5], which cannot be predicted from an initial threshold measurement during system fitting. However, routinely collected measures were nonetheless important to the follow-up model’s predictions.

In all three scenarios, and with each model type, time since blindness onset (‘SubjectTimeBlind’) proved to be highly predictive of future electrode deactivations.

## V. Discussion

The present study is a retrospective investigation of a large clinical dataset and demonstrates the untapped value in clinical recordings taken from neuroprostheses. In this work, we demonstrate the prediction of electrode deactivation and perceptual thresholds using XAI models and the insights into measurable factors influencing perceptual sensitivity that can be gleaned from these models. Automating threshold prediction using imaging and clinical data may be an important and cost-effective strategy for retinal implant calibration.

On a longitudinal dataset composed of data from 12 subjects with Argus II retinal prostheses, electrode deactivations were predicted with AUC values from 0.475 when exclusively using routine clinical measures up to 0.813 when incorporating system fitting data and 0.913 when leveraging information from previous examinations. Additionally, perceptual thresholds were predicted using routine, system fitting, and followup measurements, with associated 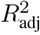 values of up to 0.765. On the one hand, these findings highlight the importance of periodical threshold measurements to continuously monitor device performance. On the other hand, in the absence of such measurements, our work demonstrates that routinely collected clinical measures and a single session of system fitting might be sufficient to inform an XAI-based threshold prediction strategy. Although the results presented in this study were based on measurements exclusively from the Argus II retinal implant, the predictors that we analyzed are likely highly relevant to a wider range of retinal prostheses.

Unfortunately, we did not have access to fundus photographs at each session of the original dataset. We therefore had to assume that the location of the array stays stable over time, which is supported by a recent study highlighting the long-term stability of the Argus II implant [14].

Consistent with previous studies [2]–[4], we found that electrode impedance is an important predictor of perceptual thresholds. In addition, our models discovered correlations with demographic factors, demonstrating that thresholds tend to increase with subject age, time since blindness onset, and time since implantation. This is not surprising, as RP is a progressive disease that may lead to worsening visual outcomes as time progresses. Interestingly, the importance of electrode-retina distance for the purpose of threshold prediction was age-dependent (Fig. 6): whereas low electrode-retina distances were associated with lower perceptual thresholds for the youngest subjects, the opposite was true for the oldest subjects in the dataset. For a wide range of subject ages (between − 1.5 and +2 in normalized age), electrode-retina distances were not predictive of perceptual thresholds.

**Fig. 6:**
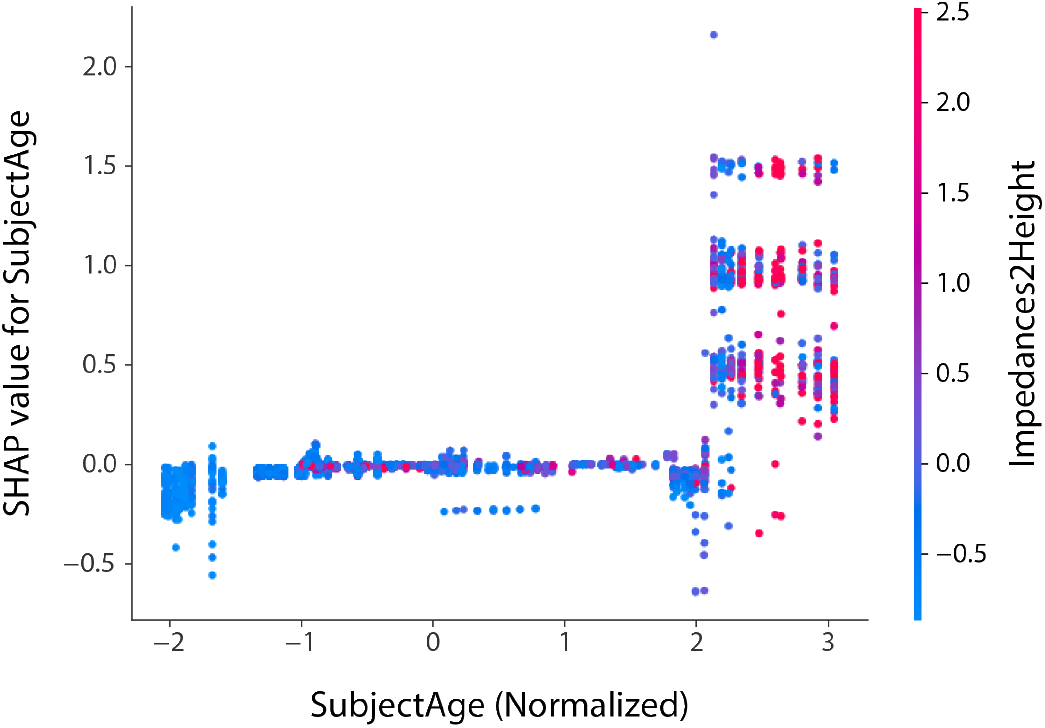
SHAP dependencies between subject age (‘SubjectAge’) and electrode-retina distance (approximated by ‘Impedances2Height’) observed in the full XGB-R model (‘Follow-Up’). Positive SHAP values indicate that the model was pushed towards predicting higher thresholds. SHAP magnitude indicates the strength of the effect. For instance, the data points on the far left of the plot (i.e., the youngest subjects in the dataset) indicate that low electrode-retina distances (blue) were correlated with lower perceptual thresholds (negative SHAP values). For subjects above normalized age +2, electrode-retina distances were strongly correlated with higher perceptual thresholds (large positive SHAP values).

Interestingly, our models also identified electrode-fovea distance (i.e., retinal eccentricity) as an important predictor of perceptual thresholds. As RP progresses from the periphery inwards, this and other neuroanatomical markers could stand in as a proxy for disease progression, which may be important for predicting visual outcomes with patient-specific computational models of prosthetic vision [15], [22], [23]. However, as our study is limited to Argus II data, future work should focus on replicating these results based on data from other (and preferably: multiple) retinal implants.

To the best of our knowledge, this work (along with our preliminary study [8]) comprises a novel systematic study on perceptual sensitivity for the field of retinal prostheses. Accurate predictive models of electrode deactivation and perceptual thresholds that rely on established and interpretable clinical measures has the potential to benefit both the retinal implant and wider neuroprosthetics communities. In the near future, such data-driven approaches could complement expert knowledge-driven interventions [24], making increasingly few assumptions about the underlying data and instead automatically inferring and exploiting relationships among the measured features, thus potentially transforming clinical practice in predicting visual outcomes.

## Data Availability

Data produced in the present work are available upon reasonable request to the authors and collaborator permissions.

## Author Contributions

M.B. and A.R. compiled the longitudinal dataset and established preliminary results. G.P., Z.H., and M.B. implemented data processing, models, and evaluation. G.P. and M.B. wrote the manuscript. All authors approved the final version of the manuscript.

## APPENDIX

### A. Supplemental Data and Figures

#### 1) Subject Demographics

Summary subject demographics are provided in Table A.1. Data reported in this table relevant to subject age and the amount of time for which the subject had been blind reflects information from the earliest recording obtained from each subject. All subject demographics were provided by, or estimated from, previous studies [3], [11], [12].

**TABLE A.1:**
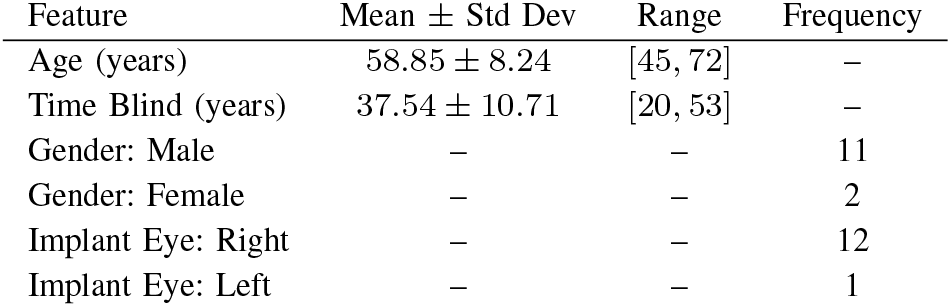
Subject Demographics

| Feature | Mean ± Std Dev | Range | Frequency |
| --- | --- | --- | --- |
| Age (years) | 58.85 ± 8.24 | [45, 72] | – |
| Time Blind (years) | 37.54 ± 10.71 | [20, 53] | – |
| Gender: Male | – | – | 11 |
| Gender: Female | – | – | 2 |
| Implant Eye: Right | – | – | 12 |
| Implant Eye: Left | – | – | 1 |

#### 2) Subject Threshold Distributions

High variance in perceptual thresholds across subjects and among the electrodes of a single implant make threshold prediction a challenging task. Kernel density estimates of perceptual thresholds for each of the twelve subjects studied in this work are provided in Fig. A.1. Perceptual threshold variance was lowest for subjects 12-005, 51-009, and 52-001. Qualitatively, higher 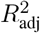 values were typically observed in LOSO regression evaluation for these subjects.

**Fig. A.1:**
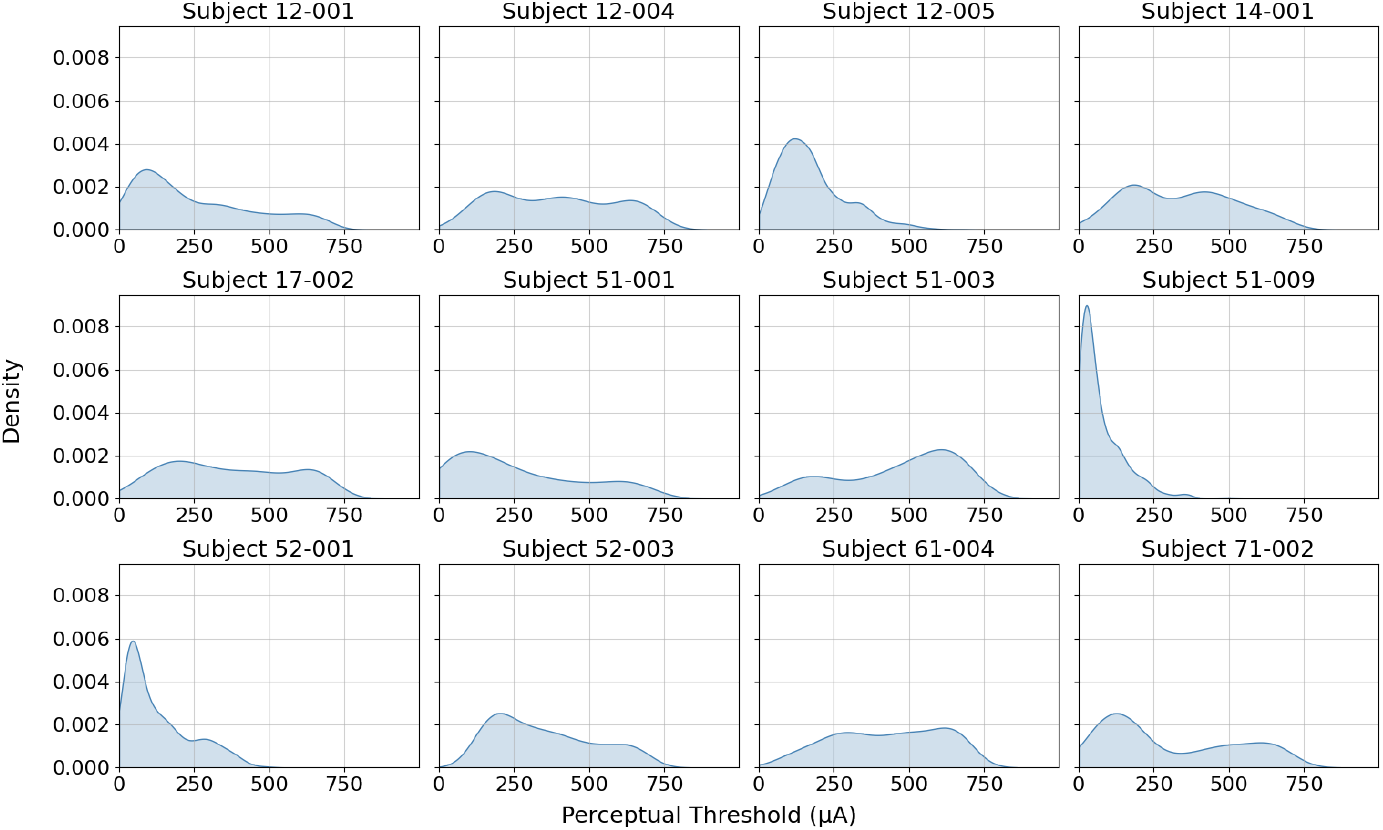
Kernel density estimates of perceptual thresholds for each subject.

#### B. Per-subject Regression Model Fit

A closer inspection of regression model outputs at the subject level reveals additional insights into EN and XGB-R model behavior when fitted to each subset of feature data. When fitted to ‘Routine’ features, the linear EN model predicted relatively consistent perceptual thresholds for nine of the twelve subjects (Fig. B.2, top), likely suggesting poor linear relationships between ‘Routine’ predictors and perceptual thresholds. Similar behavior can be observed from the EN models fitted to ‘System Fitting’ data (Fig. B.2, middle) and is additionally reflected in the low 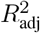 values for these models. Introducing ‘Follow-Up’ features significantly improved explained perceptual threshold variance for all subjects (Fig. B.2, bottom), and enabled 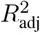 values greater than 0.7 for five of the twelve subjects. The higher variance XGB-R model also failed to capture generalizable relationships between ‘Routine’ predictors and perceptual thresholds (Fig. B.3, top), despite our finding that some of these predictors (including, but not limited to, time since blindness onset, electrode impedance, and electrode-fovea distance) are important to threshold prediction. When ‘System Fitting’ features were included, improvements in explained variance can be observed for a few of the subjects but there were still a number of subjects whose perceptual thresholds were not well modeled (Fig. B.3, middle). 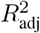 values for all subjects, with the exception of 51-001, improved when the XGB-R model was fitted with ‘Follow-Up’ features (Fig. B.3, bottom). Similar to the results observed with the EN model, highest 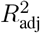 values were observed for subjects 12-001, 12-005, 52-001, and 71-002.

**Fig. B.2:**
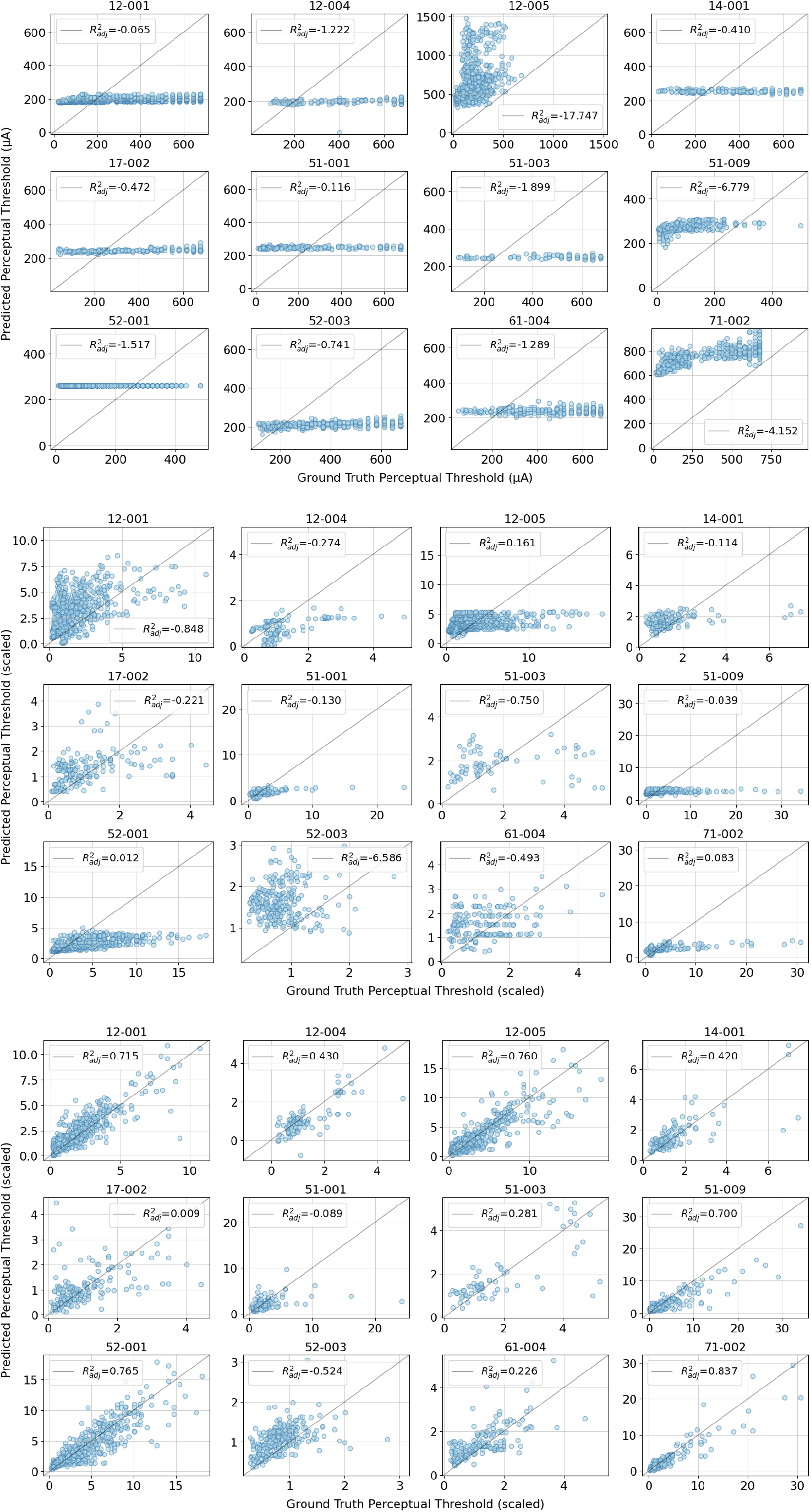
Ground truth and EN predicted perceptual thresholds based on ‘Routine’ measures (top), ‘System Fitting’ measures (middle), and ‘Follow-Up’ measures (bottom).

**Fig. B.3:**
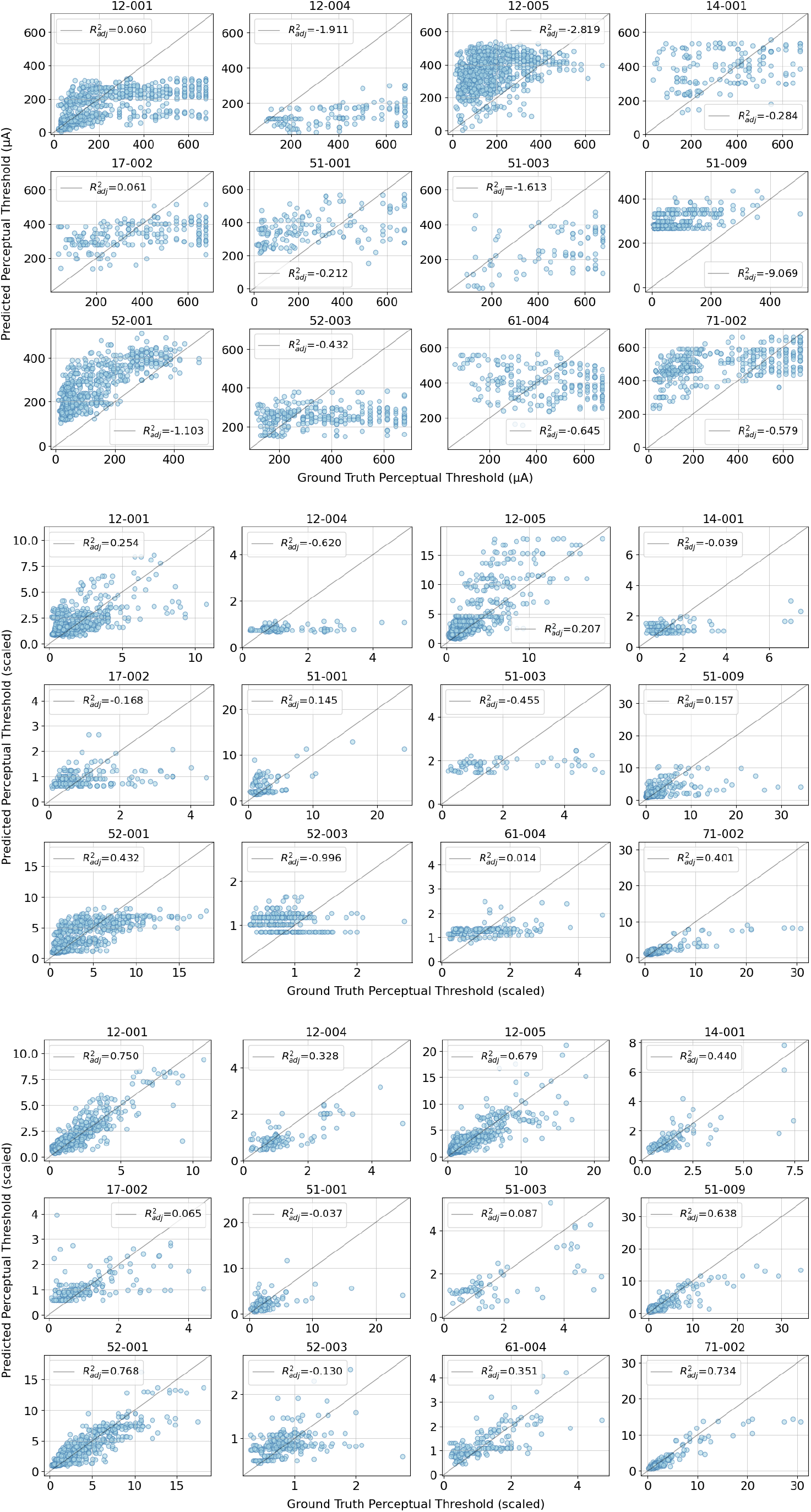
Ground truth and XGB-R predicted perceptual thresholds based on ‘Routine’ measures (top), ‘System Fitting’ measures (middle), and ‘Follow-Up’ measures (bottom).

